# Into the thirteenth Month: A Case Study on the Outbreak Analytics and Modeling the spread of SARS-CoV-2 Infection in Pune City, India

**DOI:** 10.1101/2021.06.22.21259295

**Authors:** Joy Monteiro, Bhalchandra Pujari, Sarika Maitra Bhattacharrya, Anu Raghunathan, Ashwini Keskar, Arsh Shaikh, Prasad Bogam, Shweta Kadu, Nikita Raut, Devendra Vavale, Rupa Mishra, Ajit Kembhavi, L.S. Shashidhara, Vidya Mave

## Abstract

With more than 140 million people infected globally and 3 million deaths, the COVID 19 pandemic has left a lasting impact. A modern response to a pandemic of such proportions needs to focus on exploiting all available data to inform the response in real-time and allow evidence-based decision-making. The intermittent lockdowns in the last 13 months have created economic adversity to prevent anticipated large-scale mortality and relax the lockdowns have been an attempt at recovering and balancing economic needs and public health realities. This article is a comprehensive case study of the outbreak in the city limits of Pune, Maharashtra, India, to understand the evolution of the disease and transmission dynamics starting from the first case on March 9, 2020. A unique collaborative effort between the Pune Municipal Corporation (PMC), a government entity, and the Pune knowledge Cluster (PKC) allowed us to layout a context for outbreak response and intervention. We report here how access to granular data for a metropolitan city with pockets of very high-density populations will help analyze, in real-time, the dynamics of the pandemic and forecasts for better management and control of SARS-CoV-2. Outbreak data analytics resulted in a real-time data visualization dashboard for accurate information dissemination for public access on the epidemic’s progress. As government agencies craft testing and vaccination policies and implement intervention strategies to mitigate a second wave, our case study underscores the criticality of data quality and analytics to decode community transmission of COVID-19.

## INTRODUCTION

Following a cluster of pneumonia cases recorded in Wuhan, China, in December 2019, caused by a novel coronavirus SARS-CoV-2 (1, 2), COVID-19 subsequently spread to other cities and countries (3, 4). India tops the charts of people at risk of SARS-CoV-2 infection exceeding 1.4 billion cases. The first documented case in India was from an Indian national evacuated from China on January 30, 2020. Pune, a city with about 5 million population (5), recorded its first case on March 9, 2020, with a national flying from Dubai. Severe public health measures were implemented in Pune and across India to prevent the spreading of the outbreak, including a complete lockdown starting from March 25 and dragging on for a little more than two months into the beginning of June 2020 (6). At the end of April 2021, Pune recorded a staggering >400,000 cases (7). Surveillance and contact tracing are critical components of effective public health response to COVID-19 as has been showcased worldwide, including two states of the Indian Subcontinent, Tamil Nadu and Andhra Pradesh (6).

In response to public health crises such as this pandemic, healthcare preparedness, rapid action from local authorities for procuring essential supplies, and appropriate resource allocation require projecting the trajectory of cases for coming weeks and months. Real-time advanced data analytics and computational modeling using granular data are central to such an exercise (8-11). The Pune Knowledge Cluster (PKC), a consortium of Pune-based scientists, engineers, technologists, academicians, students that believed in the power of data science analytics and modeling to curb the spread of the disease, collaborated with Pune Municipal Corporation (PMC) to provide data-driven management of the pandemic. Here, we present a perspective of the PMC-PKC collaborative efforts that include centralized data abstraction, curation, analysis, data visualization, and modeling of the early COVID-19 data to project the pandemic curve that directed policy decisions on resource procurements and allocation (12).

## ANALYTIC APPROACH

### Data Sources

Case data from the PMC included a description of reported cases gathered inline lists, i.e., flat files where each row is a case and each column a recorded variable (e.g., dates of onset and admission, gender, age, location). This data was extensively curated using a combination of manual input and state-of-the-art data science tools to fulfill the definition of ‘tidy data’ in the data science community (13). Regional level demographic information (example. age stratification, sex, residential addresses, maps of population densities) was collected independently from the smart city public data portal to delineate the underlying characteristics of the affected populations. Our primary data was obtained directly from the Pune Municipal Corporation and official press releases as part of an effort to build an epidemiological surveillance dashboard for Pune city (7). The details of the number of records and the duration for which they are available is present in **Supplementary Table 1**.

### Geocoding Methodology

Due to the unstructured nature of addresses in India, assigning an address to a geographical location is a challenging task (14). To understand the spatial evolution of the pandemic within PMC limits, we developed a machine learning model that processed addresses of each record and assigned each address to a prabhag, an administrative unit in PMC. To train the model, we used a database of 48000 addresses which experts had manually assigned in PMC to their respective prabhags. We used 80 percent of the data for training and 20 percent for validation.

We first created a database of localities around Pune, which were then assigned to their respective prabhags to eliminate the addresses’ most “noisy” parts. We first simplified and normalized the addresses using the following rules. The address was tokenized into individual words. All “small” words -- tokens of length lesser than 3, were discarded. Minimal replacement of abbreviations was done. For example: Soc -> Society, Apt -> Apartment etc. All non-alphabets are eliminated. Fuzzy matching (15) was used to replace identified tokens (1 and/or 2-grams) with a standardized form. For example, yerwada, yerawada, yerwada were all mapped to yerawada. Compound words were replaced by concatenated versions (e.g., Laxmi Nagar replaced by Laxminagar). These addresses were converted to a vector using a term frequency vectorizer. To emphasize certain unique localities within a prabhag, their frequency was multiplied by three. In the term frequency vectorization, we omit those tokens which rarely appear in the dataset. The threshold was chosen based on when the number of tokens on either side of the threshold is dramatically different. The threshold identified was 0.00003, which gives us a vocabulary of ∼5000 tokens. If the threshold was changed to 0.00004 and higher, the size of the vocabulary changes proportionally. However, if the threshold was decreased to 0.00002, the vocabulary more than doubles to ∼12000 tokens, indicating the presence of a large number of rare, “noisy” tokens.

### Classification and Validation

The resulting address vectors were used to train a variety of classifiers. On examining their performance, we observed that the multinomial Naive Bayes classifier provided better performance in certain prabhags than tree-based classifiers (Decision Tree, Random Forest, XGBoost). Thus, an ensemble architecture was chosen for the classifier. We used an ensemble of Multinomial Naive Bayes, XGBoost, and Random Forest classifiers with a Decision Tree classifier acting as a meta-classifier. With this architecture, we achieved an accuracy of 87%. Hyperparameters for Random Forest and XGBoost were chosen to reduce overfitting. The accuracy on the training set was ∼91.5%, indicating slight overfitting. We also performed 10-fold cross-validation, which showed minimal variability in the accuracy, suggesting that the model was robust.

### Outbreak Analytics: An Overview

Data type priorities were critically defined by what actionable information can be predicted for use in ground-level decision-making during the pandemic. As seen worldwide, the resultant outputs of the outbreak data analytics for Pune were limited due to operational constraints suggesting an urgent need for resources and capacities to ensure data availability and quality. The aspects of outbreak analytics (**Figure 1**) allowed a systematic understanding of the situation in Pune. ‘Outbreak analytics’ in this article refers to several tools and methods used to collect, curate, visualize, analyze, model Pune pandemic data. An integrated workflow represented describing the tools and their cross-talk are summarized below.

**Figure 1:**
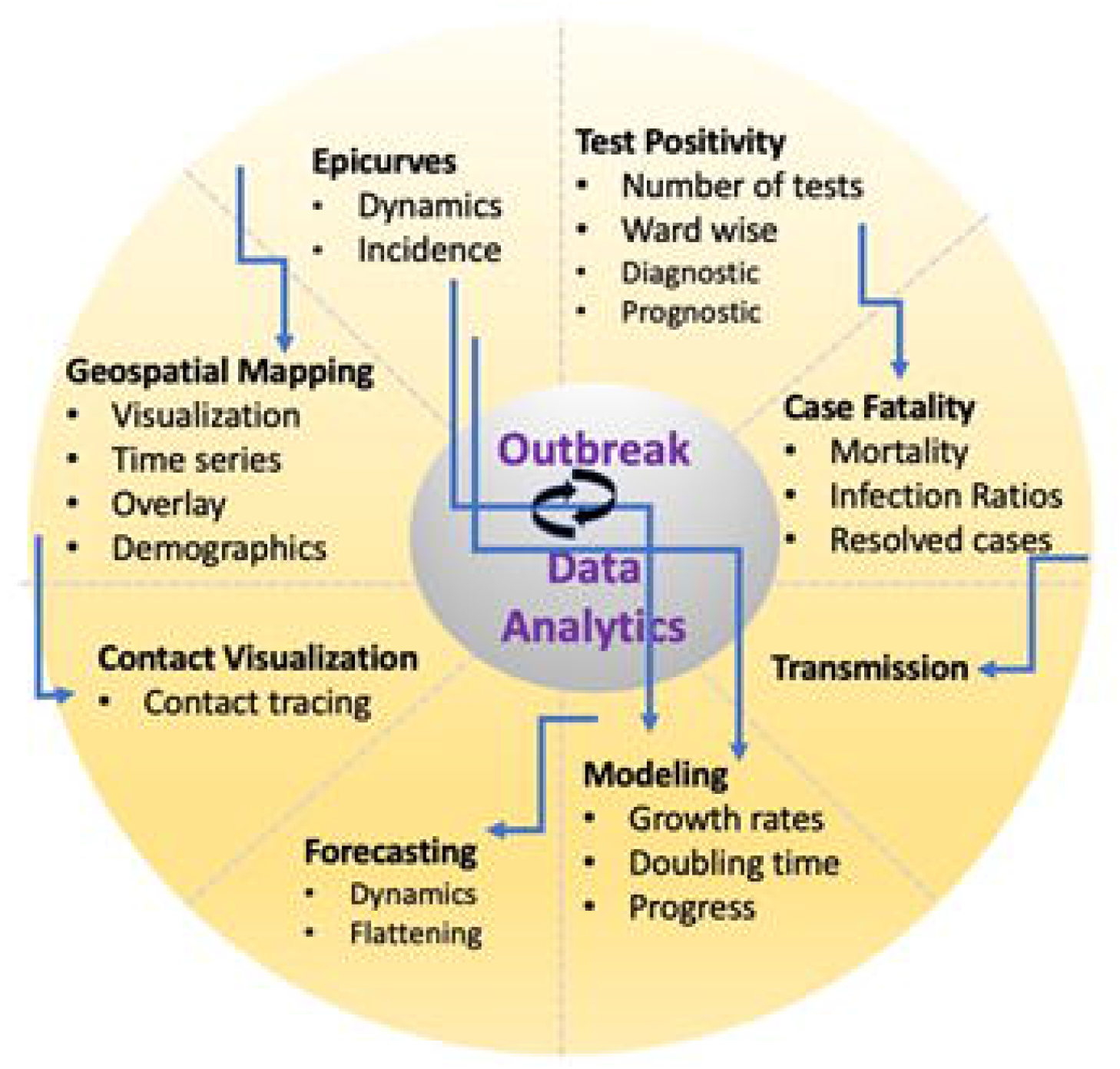
Schematic representation of tools and cross talk in the integrated pipeline for outbreak data analytics in Pune, India

### Tracking the epidemic curve

The first type of graphics needed for rapid assessment of ongoing dynamics is the epidemic curve (epi curve), which shows case incidence time series as a histogram of new-onset dates for a given time interval (16). **Figure 2a** shows the trend of cumulative cases ward-wise in Pune and their outcomes. Cumulative cases, the fraction of active cases, i.e., the ratio of active cases and total cases, is another suitable parameter to understand the state of the epidemic. In the beginning, when all cases are active, it will start from unity, and at the end of the epidemic, when all the cases are resolved, then this will become zero. Thus, this curve can vary between one and zero, and if it shows a decline, it implies that the epidemic is fading out. Note that when there are multiple waves, then this curve will be oscillatory.

**Figure 2:**
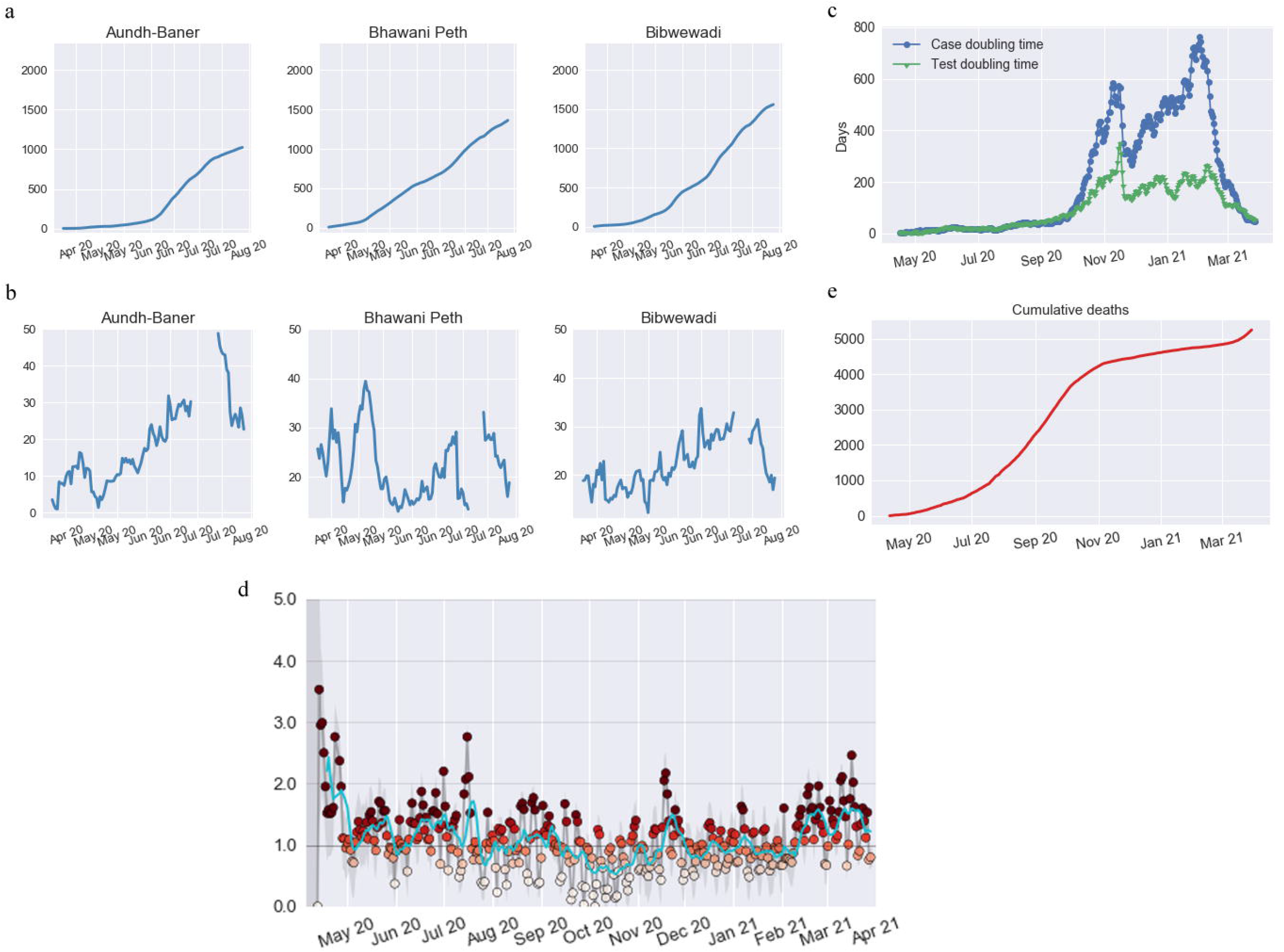
a. Presents the trend of cumulative cases ward-wise in Pune (representative wards) b. Shows the ward positivity for representative wards c. Demonstrates the test and case doubling time throughout the pandemic d. Represents the City wise R(t) e. Represents the overall cumulative deaths.

### Geo-mapping of epidemics

Maps have been at the core of infectious disease epidemiology from a very early stage. They can be typically used to visualize the distribution of disease, represent the ‘ecological niche’ of infectious diseases at large scales, assess an outbreak’s spatial dynamics, and strategize interventions (17). The changing scenario month-wise for Pune is delineated through a scale that allows understanding geographical spread **(Figure 3**). The changing geography indicates the spread of the disease from a highly infected ward with primary infection to adjacent wards.

**Figure 3:**
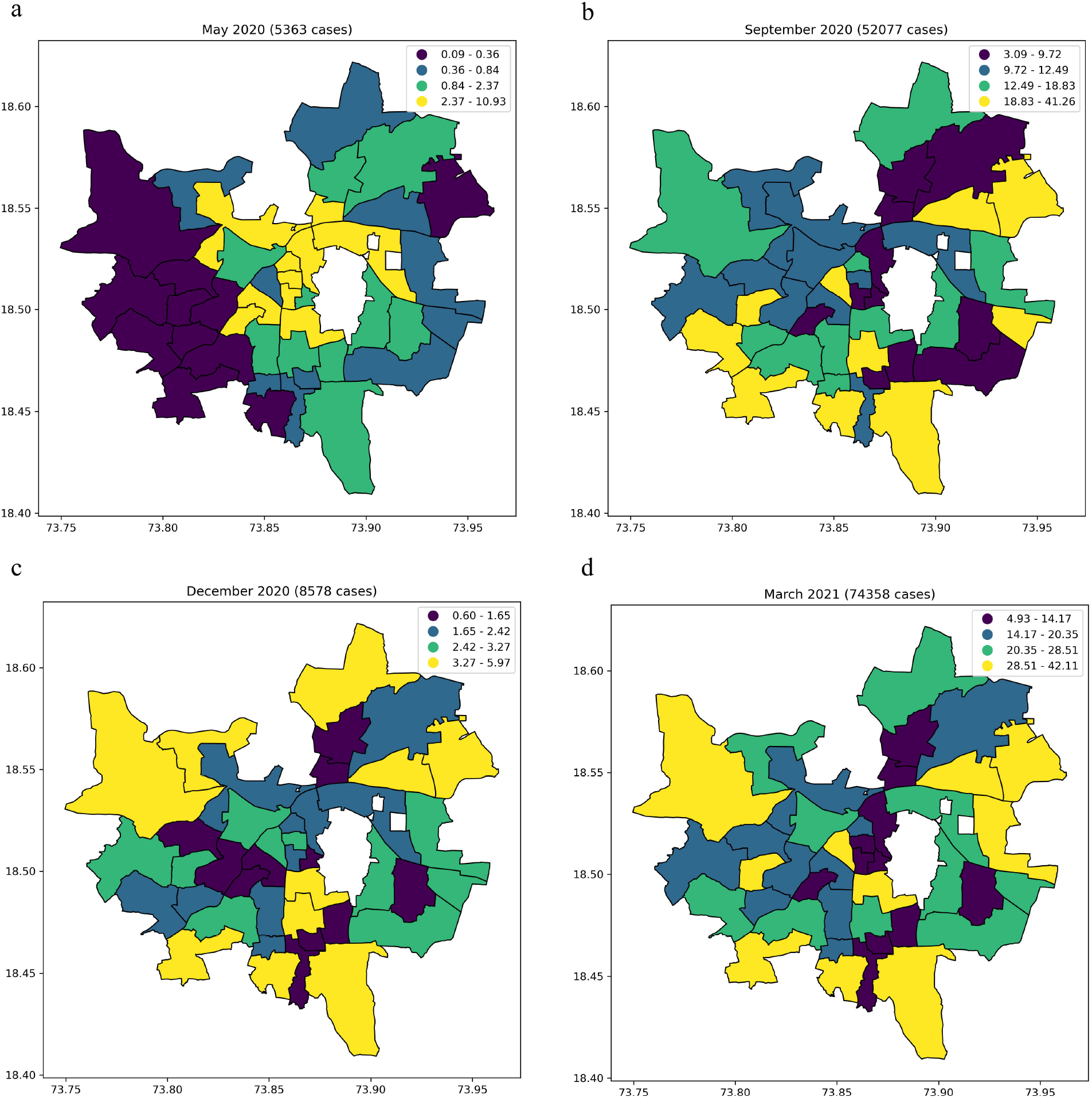
Prabhag-level incidence of COVID-19 (per 1000 persons) in Pune city for the months of May, September, December 2020, and March 2021. The incidence shows a systematic shift from the densely populated interior regions of the city towards the more sparsely populated exterior regions over time.

### Trend in test positivity and transmissibility assessment

The identification of infected cases in the population is dependent on the testing for the disease. The number of tests done in phase I of the epidemic was low and indicated that it was mainly used as a diagnostic for symptomatic people. **Figure 2b** illustrates the test positivity rates, which declined over time, indicating an increase in the number of tests. The ‘transmissibility’ of a disease can be quantified and used to refer to the rate at which new cases arise in the population, resulting either in epidemic growth or decline. Rather than an intrinsic property of a specific disease, transmissibility thus quantifies the propagation of a pathogen in a given epidemic setting and is impacted by multiple factors, including population demographics, mixing, and levels of pre-existing immunity. The first measure of transmissibility is the growth rate (r), which is estimated from a simple model where case incidence is either exponentially growing (r. 0) or declining (r, 0). Typically, r is estimated directly from epicurves using a log-linear model, where r is defined as the slope of linear regression on log-transformed incidence.

**Figure 2c** shows test and case doubling time throughout the pandemic so far. While r quantifies the speed at which a disease spreads, it does not contain information on the intervention level necessary to control the disease. This is better characterized by the reproduction number (generically noted ‘R’), which measures the average number of secondary cases caused by each primary case. Researchers typically distinguish the basic reproduction number (R0), which applies in a large, fully susceptible population, without any control measures, from the effective reproduction number (Reff), which is the number of secondary cases after accounting for behavioral changes, interventions, and declines in susceptibility. Different methodological approaches have been developed to estimate the reproduction number. To estimate Reff, we used the Bayesian approach developed by Bettencourt and Ribeiro, and modified by K. Systrom (18, 19).

R can be approximated using estimates of the growth rate r combined with knowledge of the generation time distribution. A more reliable method is to derive R(t) from compartmental models. **Figure 2d** represents the change in R0 values ward-wise over time. The prevalence of the disease can be calculated through understanding IFR as opposed to CFR. This can be calculated by serological testing to detect the %age population infected. A serosurvey with five prabhags done in the July-August 2020 time frame is discussed here in connection with the cases detected and case fatality ratios.

### Tracking Case Fatality Rates

Mortality in infectious diseases has a lag defined by the time interval between case detection and outcome. The outcome can be represented as death or recovery. **Figure 2e** shows the overall cumulative deaths. The case fatality ratios go up during phase 1 and early part of phase 2 of the epidemic, while the case fatality rates (CFR) are coming down with time. CFR calculations calculated from publicly available data face lag between the date of admission of patients and the date of death. Accounting for this lag is a major source of uncertainty in CFR calculations. Due to the availability of date of isolation in the data made available by PMC, we have calculated CFR, which reduces this uncertainty significantly, and allows for a cohort-based (i.e., all people who were isolated in the same period) calculation of CFR. Calculating CFR based on the date of isolation also allowed us to compute a “real-time” CFR which accounts for currently active cases.

### Dashboard and real-time visualization of data

In order to disseminate information about the pandemic state, an online dashboard was created and operated by the team depicting the trajectories of the epidemic (7). Dashboard depicts various quantities and figures developed by the team members such as daily and active number of cases as well as tests, real-time effective reproduction rate (*R(t)*), case doubling time, test doubling time, active fractions, cumulative test positivity, CFR and the forecasting. Such quantities were made available for the entire city; whenever possible, more granular data based on wards and sub-wards (Prabhags) was also shown. An example plot of the dashboard is shown in **Figure 4**.

**Figure 4:**
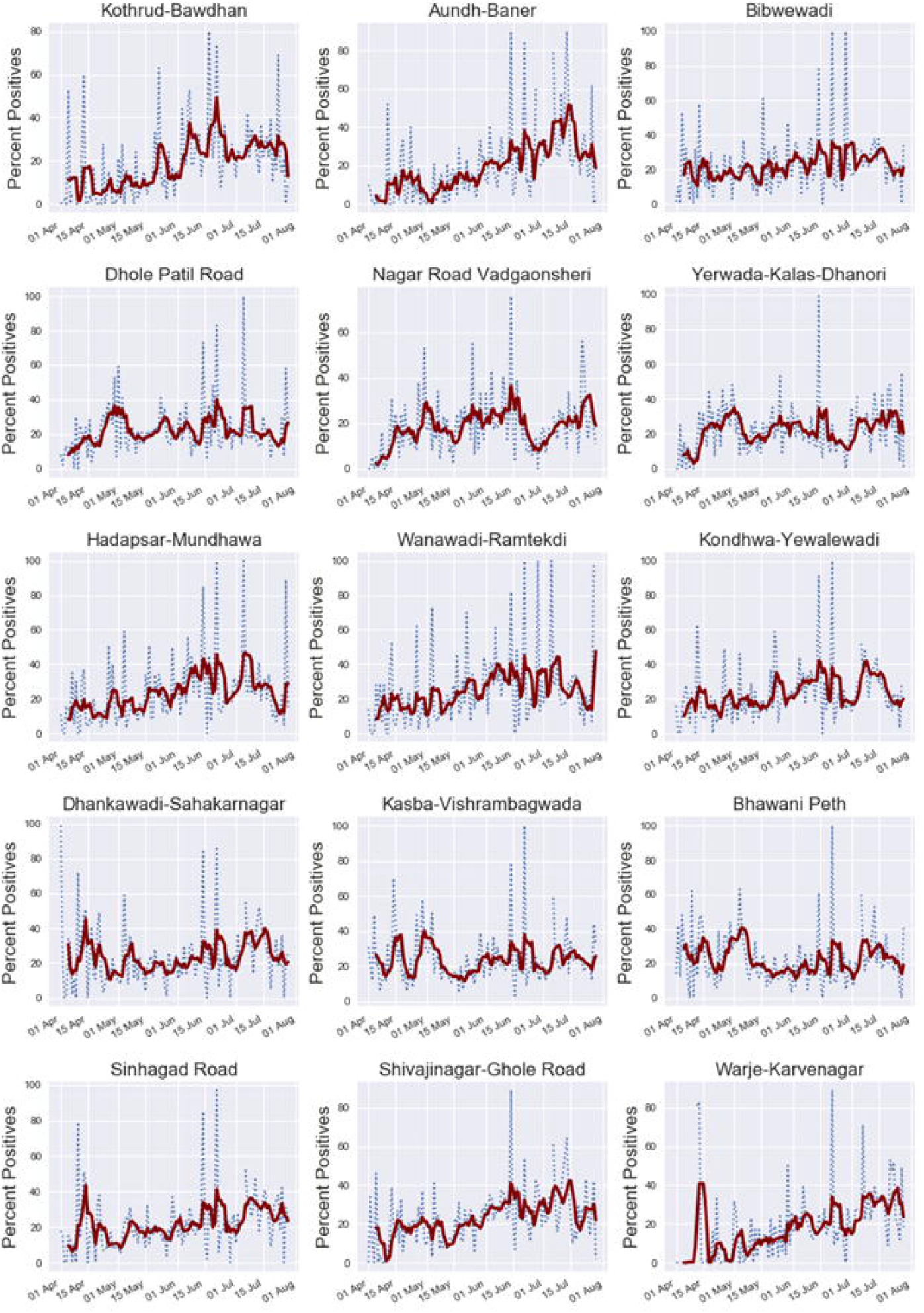
Dashboard - Example plot for Prabhag-wise cases per 1000 population

### Forecasting

Forecasting plays a vital role in policymaking by offering reasonable estimates of the upcoming burden on health infrastructure. We used the state-of-the-art INDSCI-SIM model, an extended SEIR model (20, 21) that employs a nine-compartmental description of an epidemic, tailored specifically to COVID-19. Because INDSCI-SIM supports various non-pharmaceutical interventions, one can utilize the model to incorporate the effects of multiple measures such as lockdown, adherence to masks, social distancing, and quarantining. For forecasting, we chose the number of patients in critical condition for two primary reasons: 1) this data is less prone to errors of underreporting as hospitals typically test all the critical care patients for COVID-19 and 2) this number directly affects the resource requirements to be made by the municipal corporation. Our first forecast was made in June 2020, which was subsequently updated after the second lockdown in July. The resulting forecast was made available on an online dashboard until the end of the year. The reported cases were added every day as shown in **Figure 5** that shows a worst-case and a best-case scenario.

**Figure 5:**
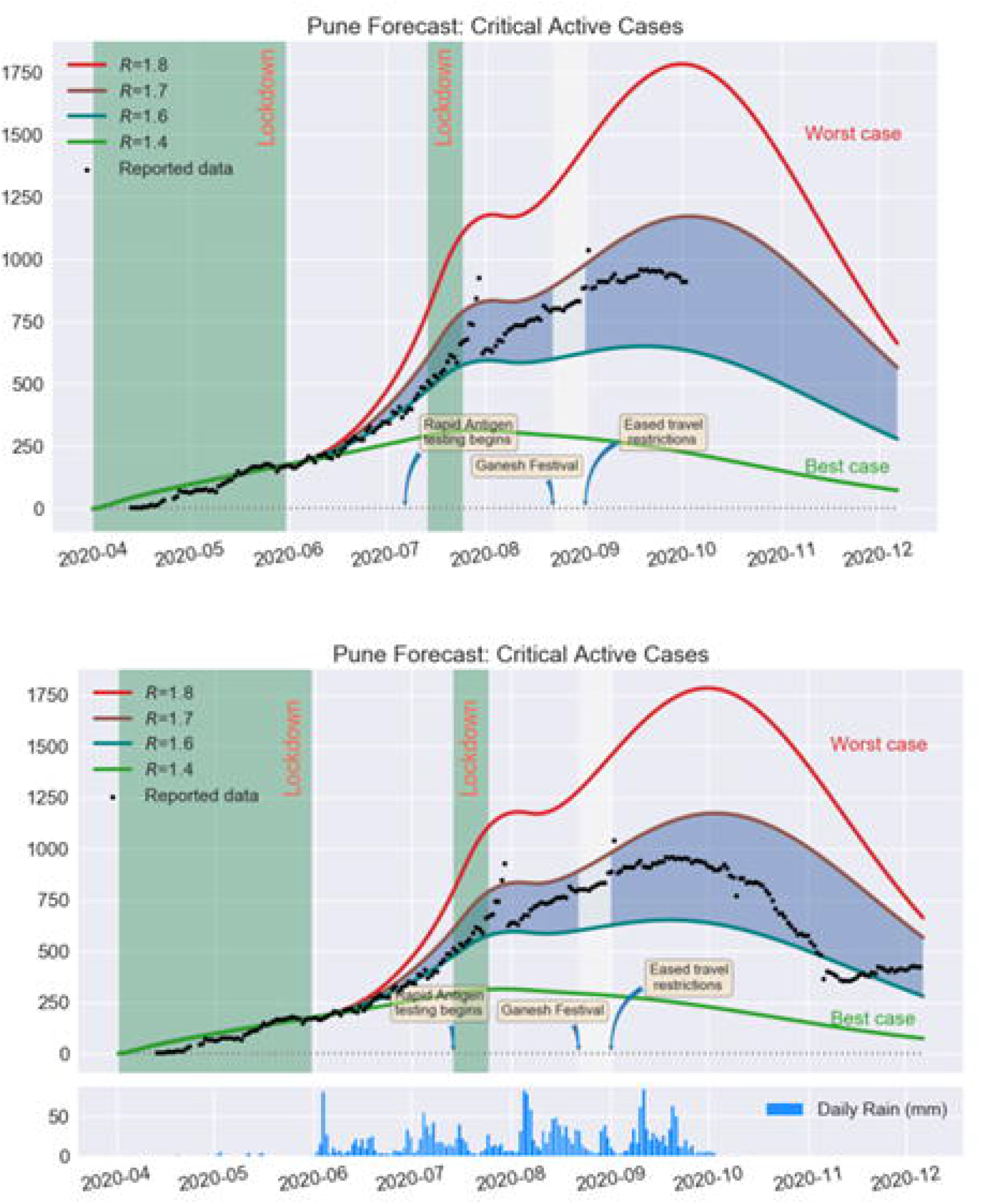
Modelling/Forecasting - Shows a worst case, a best-case scenario along with the likely path the data may take (region shaded with blue).

### How much should we test?

The way to control the infection is to increase the number of tests. However, can we quantify how much should be good enough? We proposed that if the time doubling value of the test is lower than that of the time doubling the value of infection, it is possible to test a larger population beyond just symptomatic people and their contacts. In the worst-case scenario, the two-time doubling values should be at least the same. This was done when PMC ramped up their tests from about 700-800 per day at the end of May to approximately 2500 in mid-June. Was that enough?

Like the time doubling of cases, we can define the time doubling of tests as

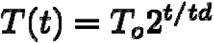

To -- the cumulative number of tests on June 16 = 76206.

td-- time doubling value

T(t)-- Cumulative number of tests on a day starting from June 16.

We projected values of tests required to bring down the test doubling time (td). Note that the case doubling time during that period was varying between 22-28 days. The projected tests required for keeping the td value at a certain number over the next 19 days are given in the table. These numbers were much higher than the number of tests done per day. This projection gave an idea to PMC about the requirement of an increase in the number of tests per day (**Supplementary Table 2**).

### Estimation of undetected cases and distribution of the test using CFR

We asked here, if we have a limited number of tests how do we distribute across the wards? For this, we take the help of the ward wise CFR

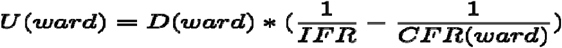

**U(ward)** = estimate of undetected cases in a ward

**D(ward)** =the deceased in that ward

**CFR(ward)** = The CFR calculated for that ward

**IFR** - can be equated to the CFR of a country or state where the epidemic is under control Total Number of undetected cases

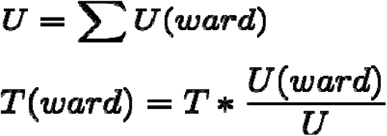

**T** --- Total per day tests for Pune

**T(ward)**--Tests per day that should be assigned to a ward

**Supplementary Table 3** shows the estimate of the ward-wise undetected cases and the number of tests required to keep the test positivity ratio - 0.2.

### Other Important Parameters

#### Test Positivity

The test positivity (TP) is defined as the ratio between the total number of positive outcomes(P) and the total number of tests (T) that may represent the testing efficacy. In an ideal case, as the number of tests is increased, this parameter should decrease or remain constant at a very low value, suggesting uniform testing and can indicate that the testing is neither targeted nor the disease is exploding undetected. High values of TP could imply the testing is targeted; that is, most tests are done only for the symptomatic patients and their contacts, enhancing the TP values artificially. A potential drawback of high TP numbers is the distinct possibility of a lower number of tests performed and, more crucially, missing out on potentially infective yet asymptomatic sections of the population, leading to a silent explosion of the disease without detection. On the other hand, a small TP value could imply higher random testing or, in the best cases, more expansive and successful testing of the population through intense contact tracing.

Given the large population and limited testing capabilities, a correct balance has to be struck between these two approaches: intensive and focused only target testing and ineffective and unplanned random testing. Thus, apart from CFR, we need to keep track of the TP. We plotted the ward-wise CFR (**Supplementary Figure 1**). Depending on the high and low values of CFR and TP, the values can be in the four-quadrant. We discuss below what each of the quadrants means and how the testing strategy can be modified.

a. Low CFR and Low TP-Low TP implies that there is not much sampling error and that the number of detected cases is close to the actual cases. When combined with low CFR values, it can be deduced that wards that come under this category have implemented good testing strategies covering most of the population. As long as the wards manage to stay in this quadrant, they can be expected to bring down the number of cases using the same strategy.
b. High CFR and High TP-This quadrant implies that testing is too targeted, and thus the sampling efficiency is poor. The actual number of cases is many-fold higher than the detected ones.
c. Low CFR and High TP-This combination suggests that though the testing is targeted, the number of actual cases is likely to be close to the number of detected cases. The wards in this quadrant might have mixed features in terms of rate and number of cases. However, the targeted testing strategy seems to be working for these wards, and as long as they can keep the CFR low, they can follow the current testing strategy. Increasing values of CFR for these wards are an early signature of community spread. The ward should immediately ramp up the number of tests and change the strategy to include random testing over a larger population.
d. High CFR and Low TPR: Though low values of TP alone can indicate a wider testing strategy spread over a larger population when considering the high values of CFR, the conclusion about the testing strategy becomes flawed. The actual cases are much larger for the ward belonging to this most undesirable quadrant than the detected ones. The wards with these values of TP and CFR need to ramp up their testing strategy by many folds and must include random testing over a larger population.

## SUMMARY

In this report, we summarize the Pune Knowledge Cluster consortium activities that included real-time data visualization for identifications of hotspots, provision of a snapshot of outbreak trends, and forecasting of the epidemic over the next several months (7, 22). Our case study on the outbreak analytics and modeling the spread of SARS-CoV-2 infection in Pune city, India, provides a feasible and scalable proof-of-concept to facilitate recommendations for public health policies to local officials and help forecast outbreaks.

## Data Availability

City level data was collated from Press Releases. More Detailed granular data was collected from the local government, Pune Municipal Corporation is not available publicly.

## ACKNOWLEDGEMENTS

We would like to thank Rubal Agrawal, Shantanu Goel and Saurabh Rao (Pune Municipal Corporation), Srinivasa Bharath Padavala (Pricewaterhouse Coopers), external advisors: Gautam Menon, Dibyendu Nandi (IISER Kolkata), Sneha Bhogale (PKC), Hemant Joshi (TCS), Rahul Jagtap (PMC), Shivik Garg, Shrutika Lokapure, Collins Assissi (IISER Pune).

## AUTHOR CONTRIBUTIONS

JM & LS conceived the study and BP, JM, SB, AR, AK1, NR, DV performed outbreak analytics and modelling. All authors BP, JM, SB, AR, AK1, AS, PB, SK, NR, DV, RM, AK2, LS & VM contributed in respective section of the manuscript. All authors declare no competing interests

## Figure List

**Supplementary Figure 1:**
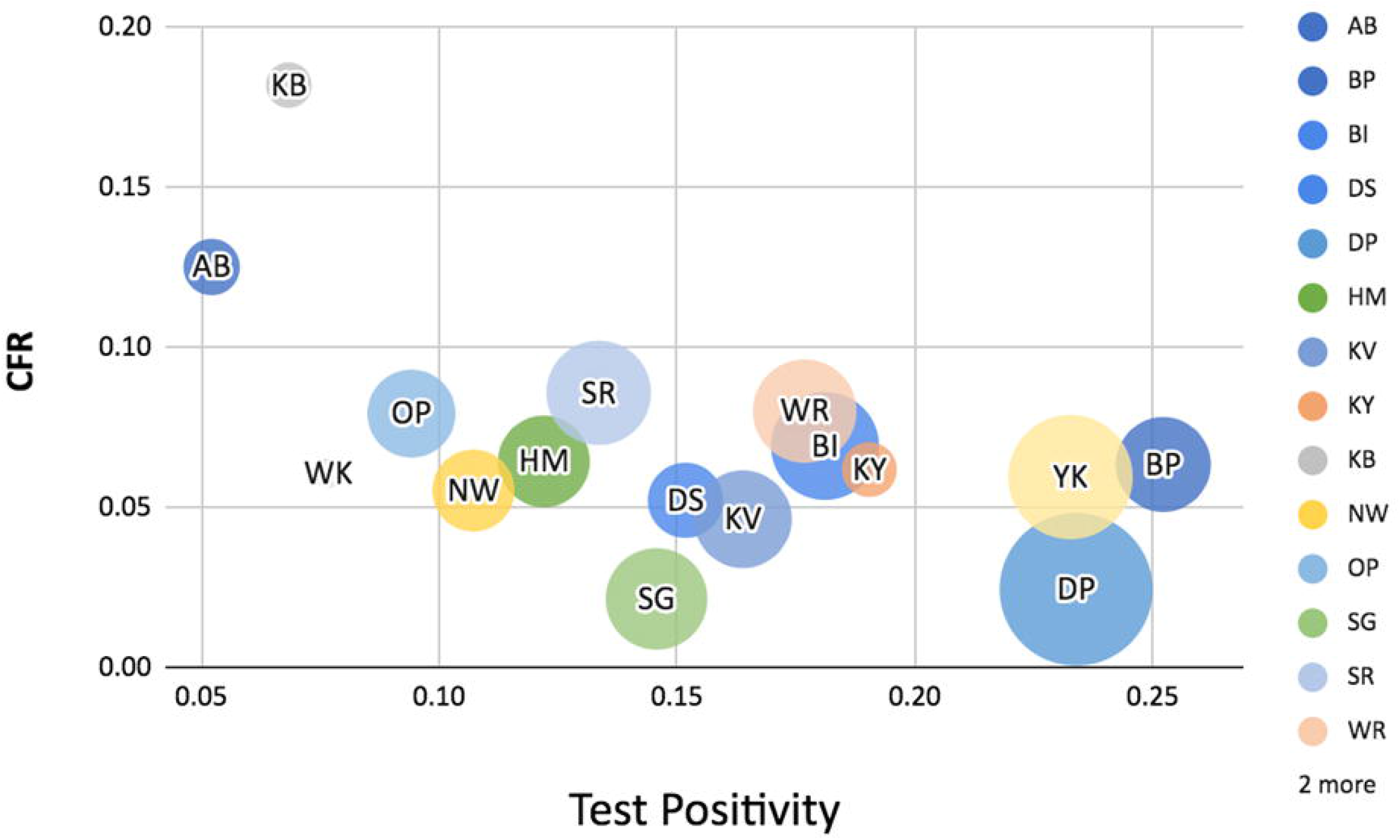
Prabhag-level Case Fatality Rates (in percent) in Pune city between February 1 - April 15, 2021. The relatively high CFR in some prabhags with low incidence suggests a higher number of undetected cases (see text for details).

**Supplementary Table 1:** Summary of characteristics available from the data sources

| Type of data | Number of Records | Total Records (from press release) | Duration | Remarks |
| --- | --- | --- | --- | --- |
| Testing | 309742 | 521249 | 2020-03-01 to 2020-09-15 | Missing records are primarily from private testing agencies |
| Patients | 114669 | 115770 | 2020-03-01 to 2020-09-11 |  |
| Mortality | 2683 | 2706 | 2020-03-01 to 2020-09-11 | Collated from official press releases, includes those who were declared COVID-positive post-mortem |
| Contact Tracing | 52292 | N/A | 2020-03-01 to 2020-06-12 |  |

**Supplementary Table 2:** Forecasting the number of SARS-CoV-2 tests for Pune city, based on th targeted time doubling ‘td’ of tests.

| Date | td = 20 | td =15 | td = 10 |
| --- | --- | --- | --- |
| 6/17/2020 | 2782 | 3775 | 5862 |
| 6/18/2020 | 2880 | 3953 | 6283 |
| 6/19/2020 | 2982 | 4140 | 6734 |
| 6/20/2020 | 3087 | 4336 | 7217 |
| 6/21/2020 | 3195 | 4541 | 7735 |
| 6/22/2020 | 3309 | 4756 | 8290 |
| 6/23/2020 | 3425 | 4981 | 8885 |
| 6/24/2020 | 3546 | 5216 | 9523 |
| 6/25/2020 | 3671 | 5463 | 10207 |
| 6/26/2020 | 3800 | 5721 | 10939 |
| 6/27/2020 | 3934 | 5992 | 11724 |
| 6/28/2020 | 4073 | 6275 | 12566 |
| 6/29/2020 | 4216 | 6572 | 13468 |
| 6/30/2020 | 4365 | 6883 | 14434 |
| 7/1/2020 | 4519 | 7208 | 15470 |
| 7/2/2020 | 4679 | 7549 | 16581 |
| 7/3/2020 | 4844 | 7906 | 17771 |
| 7/4/2020 | 5015 | 8280 | 19046 |
| 7/5/2020 | 5192 | 8672 | 20413 |

**Supplementary Table 3:**
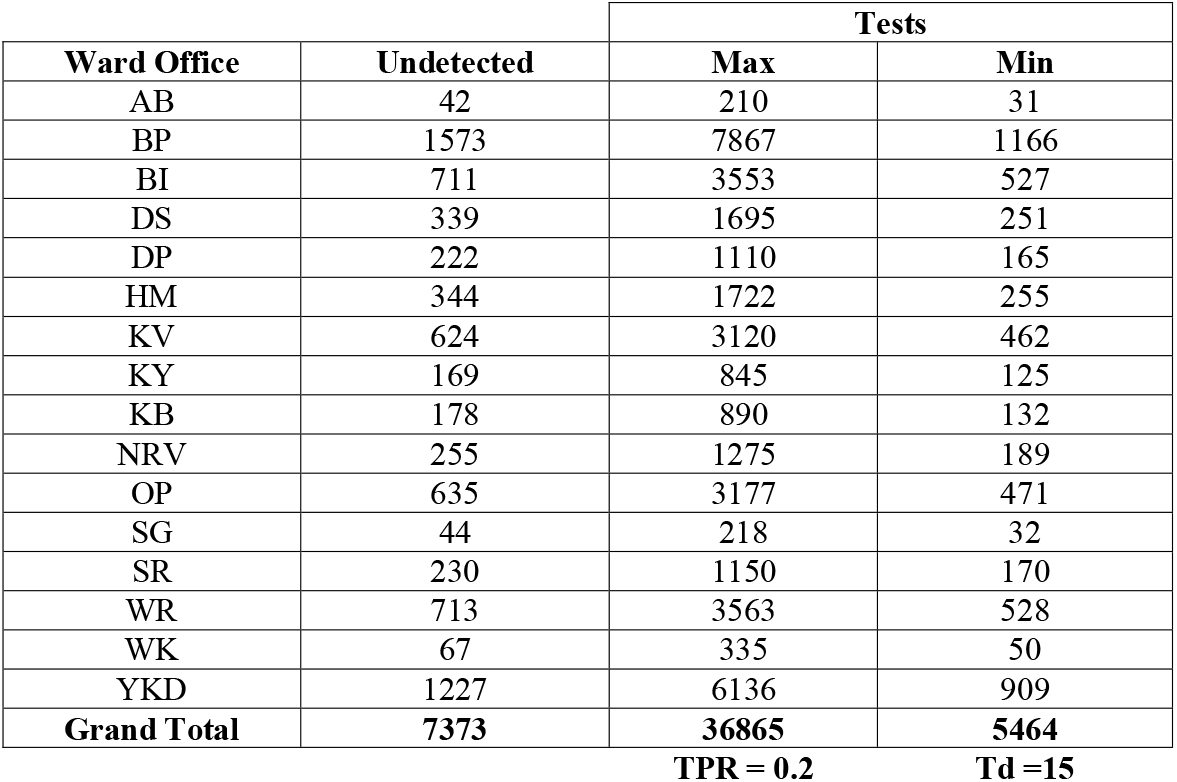
Estimation of undetected cases in the different ward offices U(ward) in Pune city based on the ward-wise CFR values and assuming the IFR to be 1%. The distribution of tests acros the wards using the information of the Total vs. ward wise undetected cases 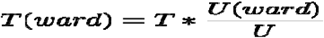

